# The Impact of Malnutrition on Host Responses to Severe Infection in Adults: A Multicenter Analysis from Uganda

**DOI:** 10.64898/2026.04.20.26351315

**Authors:** Gabriel Conte Cortez Martins, Julius J. Lutwama, Nicholas Owor, Joyce Namulondo, Jesse E. Ross, Xuan Lu, Ignatius Asasira, Tonny Kiyingi, Christopher Nsereko, John Bosco Nsubuga, Joseph Shinyale, Moses Kiwubeyi, Rittah Nankwanga, Kai Nie, Steven J. Reynolds, John Kayiwa, Seunghee Kim-Schulze, Barnabas Bakamutumaho, Matthew J. Cummings

## Abstract

**Objective:** Studies of nutritional status and host responses during severe and critical illness have focused predominantly on obesity; in contrast, the relationship between undernutrition, host responses, and clinical outcomes in adults hospitalized with severe infection remains poorly defined. We sought to determine whether severe undernutrition is associated with distinct host responses and clinical outcomes in adults hospitalized with severe infection.

**Design:** Prospective cohort study.

**Setting:** Two public referral hospitals in Uganda.

**Patients:** Non-pregnant adults (≥18 yr) hospitalized with severe, undifferentiated infection.

**Interventions:** None.

**Measurements and Main Results:** We analyzed clinical data and serum Olink proteomic data from 432 participants (median age, 45 yr [IQR, 31–57 yr]; 44% male). Overall, 213 participants (49%) met prespecified criteria for undernutrition, including 52 (12%) with severe undernutrition. Clinically, severe undernutrition was associated with HIV coinfection, microbiologically diagnosed tuberculosis, greater physiological instability, and higher mortality. After adjustment for age, sex, illness duration, study site, and HIV, malaria, and tuberculosis coinfection, severe undernutrition was associated with higher expression of proteins involved in pro-inflammatory immune signaling, endothelial and vascular remodeling, hypoxia and oxidative stress responses, and extracellular matrix remodeling, together with lower expression of proteins linked to growth signaling, anticoagulant regulation, and lipid homeostasis.

**Conclusions:** Severe undernutrition is associated with a distinct high-risk clinical phenotype and biologic signature in adults hospitalized with severe infection. These findings suggest that undernutrition may potentiate key domains of sepsis pathobiology, with implications for strengthening nutritional support and informing host-directed treatment strategies in low- and middle-income countries where malnutrition is common.

**Key Points:** *Question:* How does undernutrition influence immune, metabolic, and endothelial responses to severe infection in adults?

*Findings:* In this multicenter cohort study of 432 adults hospitalized with severe infection in Uganda, severe undernutrition was associated with greater physiologic instability, higher mortality, and a distinct proteomic host-response profile. Adults with severe undernutrition exhibited a proteomic signature characterized by pro-inflammatory immune signaling, endothelial and extracellular matrix remodeling, and hypoxia and oxidative stress responses, together with lower expression of proteins involved in growth signaling, anticoagulant regulation, and lipid homeostasis.

*Meaning:* Severe undernutrition is associated with a distinct high-risk clinical and biologic phenotype during severe infection, with implications for nutritional support, risk stratification, and host-directed therapeutic strategies, particularly in low- and middle-income countries.

## Introduction

The global burden of sepsis is concentrated in low- and middle-income countries (LMICs), yet most studies of sepsis pathobiology have been conducted in high-income countries (HICs) (1,2). This limitation is particularly relevant in sub-Saharan Africa, where severe infection arises within a distinct host-pathogen landscape shaped by high burdens of HIV, tuberculosis (TB), malaria, and malnutrition (3,4). In HIC populations, the relationship between nutritional status and severe infection has been studied predominantly through the lens of obesity, whereas the effects of undernutrition on severe infection outcomes and host-response biology remain poorly defined, particularly among adults in LMICs (5,6). In this setting, the biologic effects of undernutrition may interact with acute and chronic infection to promote host-response dysregulation. Acute and chronic infection can induce inflammatory and catabolic stress, whereas undernutrition may further perturb the host response through enteropathy with microbial translocation, oxidative stress, progressive tissue catabolism, and impaired immune priming and T cell effector function (6). Although interactions among undernutrition, host-response dysregulation, and clinical outcomes have been described in children with severe infection in LMICs, corresponding data in adults remain limited (7). To address this gap, we analyzed clinical and proteomic data from a prospective multicenter cohort of adults hospitalized with severe infection in Uganda across the nutritional spectrum. We hypothesized that severe undernutrition would be associated with greater clinical severity and worse outcomes and, independent of other host factors and high-burden co-infections, with a distinct host-response profile characterized by pro-inflammatory immune activation, endothelial dysfunction, and tissue injury.

## Materials and Methods

We analyzed data from the RESERVE-U-2 cohort, a prospective study conducted at two public hospitals in Uganda: Entebbe Regional Referral Hospital (urban) and Tororo General Hospital (rural). Details of the RESERVE-U-2 study have been published (8). Briefly, non-pregnant adults aged ≥18 years were eligible if they had evidence of infection, defined by a history of fever or axillary temperature of at least 37.5°C, and illness severe enough to require hospital admission. Participants underwent standardized clinical assessments, including testing for HIV, malaria, influenza, and SARS-CoV-2 (19). For people living with HIV (PLWH), additional diagnostics included rapid testing for TB and cryptococcosis and quantification of HIV-1 viral load. As in prior sepsis studies from LMICs, nutritional status was assessed using mid-upper arm circumference (MUAC), as most patients were not ambulatory and bed weights were not routinely available (9). Using MUAC, we classified nutritional status using thresholds informed by large-scale, multinational analyses (10,11). Severe undernutrition was defined by sex-specific MUAC cutoffs of <20cm in men and <19cm in women, corresponding to a BMI of 13 kg/m^2^ (11). Mild-to-moderate undernutrition was defined as MUAC <24cm, corresponding to a BMI of 18.5 kg/m^2^, among participants who did not meet criteria for severe undernutrition (10). Participants with MUAC ≥24cm were classified as not meeting prespecified criteria for undernutrition.

From cryopreserved serum samples collected at RESERVE-U-2 enrollment, the Olink Target Immuno-oncology and Cardiometabolic panels were used to quantify 184 proteins implicated in innate and adaptive immunity, endothelial dysfunction, and metabolic dysregulation, as previously described (8). Protein abundance was measured via the Olink Proximity Extension Assay and expressed in log_2_-normalized protein expression (NPX) units.

Between-group clinical comparisons were performed using Chi-squared, Fisher exact, or Kruskal-Wallis H tests, as appropriate (*gtsummary* R package). Principal component analysis (PCA) was used to assess variance in the measured proteome by participant nutritional status, stratified as above. (*FactoMineR, factoextra*, and *corrplot* R packages). To identify proteins differentially expressed across nutritional strata, we performed covariate-adjusted differential expression analyses using two contrasts: (1) severe undernutrition vs. participants not meeting criteria for undernutrition, and (2) severe vs. mild-moderate undernutrition (*limma* R package). Proteins were considered differentially expressed at a Benjamini-Hochberg adjusted p-value≤0.05. Models were adjusted for age, sex, illness duration before enrollment, and HIV, malaria, or TB infection, variables that may affect nutritional status and/or host responses to severe infection; study site was additionally included as a fixed effect. To avoid collider bias, we did not adjust for physiological severity, which may be influenced by both nutritional status and dysregulated host responses.

Each enrolled participant or their surrogate provided written informed consent. The study protocol (“Clinical and molecular epidemiology of severe acute febrile and respiratory illness in Uganda”) was approved by ethics committees at Columbia University (AAAR1450 on December 13^th^, 2016) and Uganda Virus Research Institute (GC/127/17/02/582 on February 3^rd^, 2017). All study procedures were followed in accordance with the ethical standards of the responsible institutional committees and the Helsinki Declaration of 1975.

## Results

After excluding two patients with missing MUAC data, we analyzed clinical and proteomic data from 432 adults (median age, 45 years [IQR, 31–57]; 44% male; 48% PLWH), of whom 49% met criteria for undernutrition, including 12% with severe undernutrition (**Table 1, Supplemental Figure 1**). Adults with severe undernutrition, who were more frequently enrolled in Entebbe, were more often male and more likely to present with functional impairment, including inability to ambulate without assistance. They also had lower body temperature, lower systolic blood pressure, lower oxygen saturation, and higher Modified Early Warning Scores (MEWS). Patients with severe undernutrition were more likely to be PLWH, have non-suppressed HIV-1 viral loads, and have microbiologically diagnosed TB. Mortality tracked nutritional strata, with the highest mortality observed among participants with severe undernutrition.

**Table 1:** Patient characteristics stratified by nutritional status.

| Characteristic<sup>a</sup> | All patients<br>N = 432 | No Malnutrition<br>N=219 | Mild-Moderate<br>Malnutrition<br>N=161 | Severe<br>Malnutrition<br>N=52 | p-value<sup>b</sup> |
| --- | --- | --- | --- | --- | --- |
| **Mid-upper arm circumference, cm** | 24 (22, 26) | 26 (24, 26) | 22 (21, 23) | 18 (17, 18) | <0.001 |
| **Study Site** |  |  |  |  |  |
| Entebbe | 145/432 (33.6%) | 52/219 (23.7%) | 61/161 (37.9%) | 32/52 (61.5%) | <0.001 |
| Tororo | 287/432 (66.4%) | 167/219 (76.3%) | 100/161 (62.1%) | 20/52 (38.5%) |  |
| **Demographics** |  |  |  |  |  |
| Age, years | 45 (31, 57) | 43 (30, 55) | 47 (34, 60) | 45 (29, 59) | 0.23 |
| Male sex | 192/432 (44.4%) | 88/219 (40.2%) | 69/161 (42.9%) | 35/52 (67.3%) | 0.002 |
| Illness duration prior to enrollment, days | 6 (4, 11) | 6 (4, 9) | 7 (4, 11) | 8 (4, 13) | 0.088 |
| **Clinical Signs** |  |  |  |  |  |
| KPS at hospital admission | 50 (40, 70) | 50 (20, 70) | 50 (40, 70) | 50 (45, 60) | 0.19 |
| Unable to ambulate without assistance | 273/432 (63.2%) | 121/219 (55.3%) | 110/161 (68.3%) | 42/52 (80.8%) | <0.001 |
| Temperature, °C | 38.4 (37.7, 39.0) | 38.6 (37.8, 39.0) | 38.1 (37.6, 38.9) | 38.0 (37.4, 38.8) | <0.001 |
| Heart rate, beats/min | 98 (80, 110) | 95 (80, 110) | 100 (81, 114) | 100 (85, 120) | 0.063 |
| Respiratory rate, breaths/min | 24 (20, 30) | 24 (20, 30) | 26 (20, 30) | 24 (20, 26) | 0.047 |
| Systolic blood pressure, mmHg | 112 (100, 121) | 118 (104, 124) | 110 (100, 121) | 103 (93, 116) | <0.001 |
| Oxygen saturation, % | 96 (94, 98) | 97 (96, 98) | 96 (94, 98) | 95 (93, 97) | 0.003 |
| Encephalopathy | 107/432 (24.8%) | 54/219 (24.7%) | 44/161 (27.3%) | 9/52 (17.3%) | 0.35 |
| **Lactate and Physiological Severity Scores** |  |  |  |  |  |
| Whole-blood lactate, mmol/L | 2.4 (1.8, 3.0) | 2.4 (1.8, 3.1) | 2.4 (1.8, 3.0) | 2.4 (1.8, 3.0) | 0.83 |
| Whole-blood lactate ≥ 4 mmol/L | 41/432 (9.5%) | 23/219 (10.5%) | 14/161 (8.7%) | 4/52 (7.7%) | 0.80 |
| qSOFA ≥ 1<sup>c</sup> | 343/432 (79.4%) | 172/219 (78.5%) | 131/161 (81.4%) | 40/52 (76.9%) | 0.71 |
| qSOFA ≥ 2<sup>c</sup> | 144/432 (33.3%) | 66/219 (30.1%) | 59/161 (36.6%) | 19/52 (36.5%) | 0.36 |
| Modified Early Warning Score | 3 (2, 5) | 3 (2, 4) | 4 (3, 5) | 4 (3, 5) | 0.016 |
| Universal Vital Assessment score | 2 (1, 4) | 2 (0, 4) | 2 (1, 5) | 2 (1, 5) | 0.063 |
| **HIV status** |  |  |  |  |  |
| Person living with HIV (PLWH) | 204/426 (47.9%) | 92/216 (42.6%) | 84/159 (52.8%) | 28/51 (54.9%) | 0.083 |
| WHO HIV clinical stage 3 or 4 | 162/204 (79.4%) | 69/92 (75.0%) | 69/84 (82.1%) | 24/28 (85.7%) | 0.34 |
| Receiving ART prior to hospitalization | 158/204 (77.5%) | 74/92 (80.4%) | 64/84 (76.2%) | 20/28 (71.4%) | 0.57 |
| CD4 count, cells/mm3 | 314 (250, 500) | 350 (300, 500) | 307 (250, 551) | 350 (225, 500) | 0.62 |
| HIV-1 viral suppression | 127/204 (62.3%) | 66/92 (71.7%) | 45/84 (53.6%) | 16/28 (57.1%) | 0.038 |
| **Pathogen Diagnostics** |  |  |  |  |  |
| Malaria RDT positive | 133/432 (30.8%) | 66/219 (30.1%) | 53/161 (32.9%) | 14/52 (26.9%) | 0.69 |
| Microbiological TB positive<sup>d</sup> | 87/432 (20.1%) | 33/219 (15.1%) | 37/161 (23.0%) | 17/52 (32.7%) | 0.009 |
| Urine TB-LAM positive if PLWH | 69/196 (35.2%) | 28/88 (31.8%) | 28/82 (34.1%) | 13/26 (50.0%) | 0.23 |
| Cryptococcal ag. positive if PLWH | 16/191 (8.4%) | 5/90 (5.6%) | 9/76 (11.8%) | 2/25 (8.0%) | 0.36 |
| Influenza PCR positive | 19/420 (4.5%) | 11/214 (5.1%) | 7/156 (4.5%) | 1/50 (2.0%) | 0.76 |
| SARS-CoV-2 PCR positive | 21/420 (5.0%) | 6/214 (2.8%) | 11/156 (7.1%) | 4/50 (8.0%) | 0.084 |
| **In-hospital Management** |  |  |  |  |  |
| Received oxygen therapy | 117/432 (27.1%) | 50/219 (22.8%) | 50/161 (31.1%) | 17/52 (32.7%) | 0.13 |
| Received intravenous fluids | 335/432 (77.5%) | 173/219 (79.0%) | 120/161 (74.5%) | 42/52 (80.8%) | 0.49 |
| Received anti-malarial agent(s) | 168/432 (38.9%) | 90/219 (41.1%) | 59/161 (36.6%) | 19/52 (36.5%) | 0.63 |
| Received anti-bacterial agent(s) | 389/432 (90.0%) | 200/219 (91.3%) | 142/161 (88.2%) | 47/52 (90.4%) | 0.60 |
| Received anti-TB agent(s) | 114/432 (26.4%) | 42/219 (19.2%) | 52/161 (32.3%) | 20/52 (38.5%) | 0.002 |
| Received red blood cell transfusion | 20/432 (4.6%) | 5/219 (2.3%) | 11/161 (6.8%) | 4/52 (7.7%) | 0.040 |
| **Hospital outcomes** |  |  |  |  |  |
| Death | 23/432 (5.3%) | 8/219 (3.7%) | 10/161 (6.2%) | 5/52 (9.6%) | 0.056<sup>e</sup> |
| Discharged alive | 391/432 (90.5%) | 206/219 (94.1%) | 142/161 (88.2%) | 43/52 (82.7%) |  |
| Referred/Transferred to other facility | 18/432 (4.2%) | 5/219 (2.3%) | 9/161 (5.6%) | 4/52 (7.7%) |  |
| KPS at discharge or transfer | 70 (70, 90), 396 | 70 (70, 90), 204 | 70 (70, 90), 146 | 70 (70, 80), 46 | 0.46 |
| **30-day Outcomes** |  |  |  |  |  |
| Death at 30 days | 85/424 (20.0%) | 38/218 (17.4%) | 33/156 (21.2%) | 14/50 (28.0%) | 0.22 |
| KPS at 30 days | 80 (70, 90), 337 | 80 (60, 90), 178 | 80 (70, 90), 123 | 80 (70, 90), 36 | 0.86 |
| **60-day Outcomes** |  |  |  |  |  |
| Death at 60 days | 104/421 (24.7%) | 46/218 (21.1%) | 42/153 (27.5%) | 16/50 (32.0%) | 0.17 |
| KPS at 60 days | 80 (70, 100), 314 | 80 (70, 90), 169 | 80 (70, 100), 111 | 85 (70, 100), 34 | 0.57 |

**Abbreviations:** Karnofsky Performance Status, qSOFA: quick Sepsis-related Organ Failure Assessment, PLWH: person living with HIV; WHO: World Health Organization, RDT: rapid diagnostic test, TB: tuberculosis, LAM: lipoarabinomannan, PCR: polymerase chain reaction
**Legend:** <sup>a</sup>Dichotomous and categorical variables summarized as counts (proportions), continuous variables summarized as medians (interquartile ranges), n if fewer patients with available data than the total number of patients in the cohort; <sup>b</sup>Result of Kruskal-Wallis H, Chi-squared, or Fisher exact test, as appropriate; <sup>c</sup>Systolic blood pressure $\leq 100$ mmHg, respiratory rate $\geq 22$ breaths/min, and altered mental status, latter defined using AVPU scale; <sup>d</sup>Positive result by sputum Xpert Ultra or urine TB-LAM. <sup>e</sup>P-value reflects chi-squared test of the overall distribution of mutually exclusive hospital outcomes (death, discharged alive, referred/transferred) across nutritional strata.

We analyzed expression of 180 proteins after excluding four with high proportions of values below the estimated limits of detection for their respective Olink panels. PCA of the measured proteome showed substantial overlap across nutritional strata, although participants with severe undernutrition demonstrated modest separation from the other groups (**Figure 1C**). Covariate-adjusted differential expression analyses identified a core proteomic signature of severe undernutrition across both contrasts examined, indicating that severe undernutrition was distinct from both mild-moderate undernutrition and from participants not meeting criteria for undernutrition (**Figures 1A-1B**). Across both comparisons, severe undernutrition was associated with higher expression of proteins involved in pro-inflammatory immune signaling, including CXCL12, which regulates leukocyte trafficking; IL-15, which promotes natural killer cell activation; OSMR, which mediates IL-6 family signaling; and TNFRSF12A and TNFRSF21, TNF receptor superfamily members linked to inflammatory pathway activation, tissue repair, and cell death. Severe undernutrition was also associated with higher expression of proteins involved in endothelial remodeling, including the lymphatic marker LYVE1, the angiogenic mediators NRP1 and FGF2, and GAS6, which participates in thromboinflammatory signaling. Increased expression of CAIX and SOD1 further suggested hypoxia-related and oxidative stress responses in severe undernutrition, whereas increased DCN and EFEMP1 suggested extracellular matrix remodeling and tissue injury. In contrast, severe undernutrition was associated with lower expression of IGFBP3, a mediator of IGF-related growth signaling; SERPINA5, an anticoagulant serine protease inhibitor; and SAA4, an HDL-associated protein involved in lipid homeostasis.

**Figure 1:**
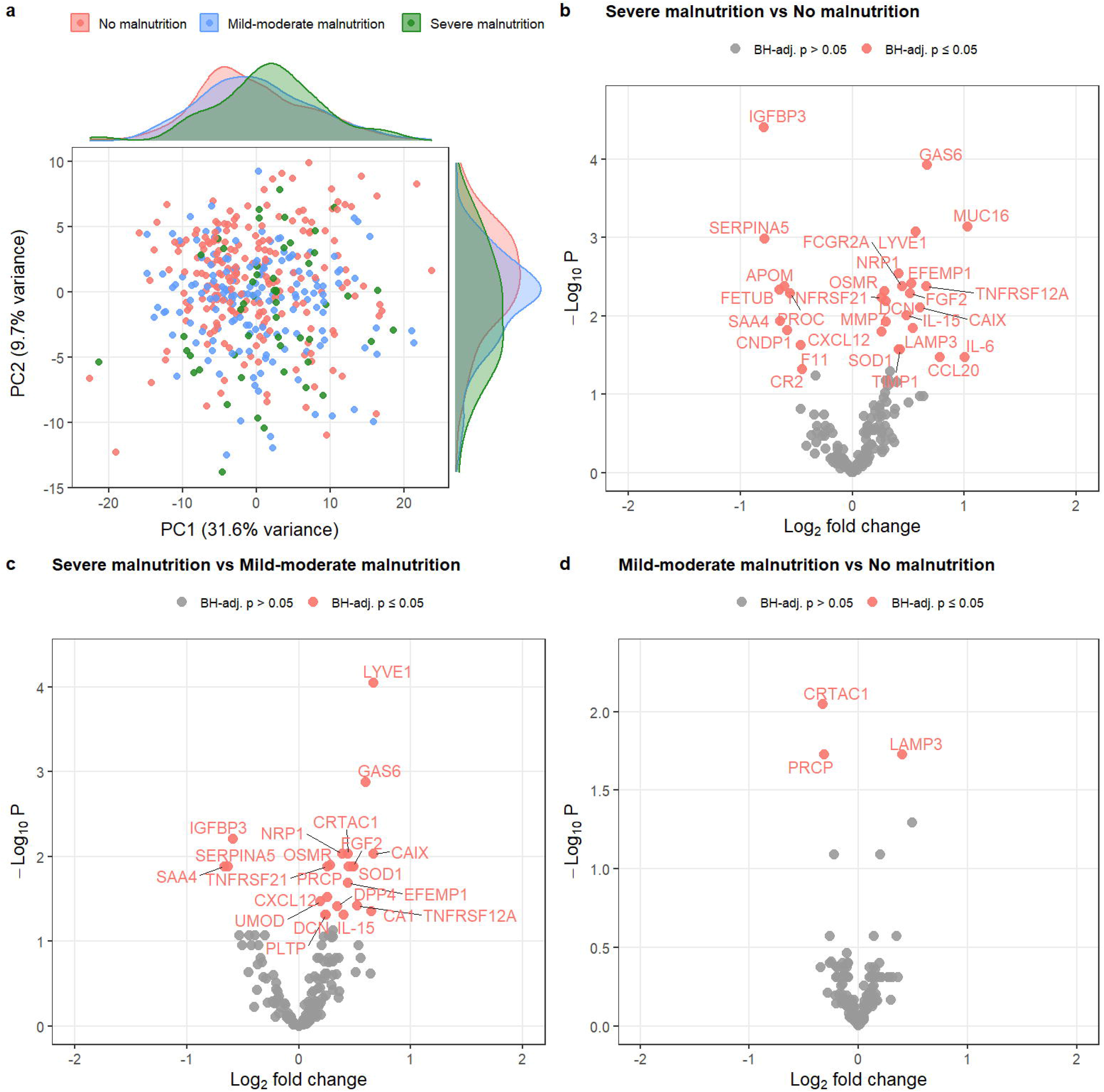
Proteomic host responses stratified by nutritional status in the RESERVE-U-2 cohort. **(a)** First two principal components of proteomic variance stratified by nutritional status; dots represent individual patients and density plots reflect individual patient distributions across principal components 1 and 2 (N=432). Volcano plots of covariate-adjusted differential protein expression in patients with **(b)** severe malnutrition vs. no malnutrition, **(c)** severe malnutrition vs. mild-moderate malnutrition, and **(d)** mild-moderate malnutrition vs. no malnutrition (N=426 across comparisons; 6 patients with missing covariate data excluded). All comparisons adjusted for age, sex, illness duration prior to enrollment, study site (as a fixed effect) and HIV, malaria, and tuberculosis co-infection. Red points indicate proteins differentially expressed at Benjamini-Hochberg-adjusted p-value ≤0.05.

## Discussion

In this prospective, multicenter cohort of adults hospitalized with severe infection in Uganda, severe undernutrition was associated with greater physiologic instability, higher mortality, and a distinct proteomic profile. These findings extend prior work from pediatric populations by showing that, in adults, severe undernutrition is associated not only with worse clinical outcomes but also with a distinctive host-response signature characterized by pro-inflammatory immune activation, microvascular dysfunction, hypoxia-related signaling, and tissue remodeling (6,7).

Our clinical findings reinforce observations from pediatric populations showing that severe undernutrition identifies a particularly high-risk subgroup of adults with severe infection, including those with more severe physiological derangement and higher mortality risk (12). Severe undernutrition was also associated with HIV co-infection with lack of viral suppression, and microbiologically diagnosed TB. Together, these findings suggest that severe undernutrition contributes to a clinically and microbiologically distinct phenotype of severe infection in sub-Saharan Africa, one shaped by both acute physiological instability and high-burden co-infections (13).

Biologically, the proteomic signature of severe undernutrition suggested multisystem host-response dysregulation rather than isolated immune activation alone. The concurrent upregulation of inflammatory, endothelial, hypoxic, oxidative stress, and matrix-remodeling pathways is consistent with a state of ongoing tissue stress, vascular injury, and maladaptive repair, whereas the downregulation of proteins linked to anabolic, anticoagulant, and lipid homeostatic pathways suggests impaired physiological reserve. Collectively, these findings support the interpretation that severe undernutrition is associated with diminished host resilience during severe infection and may potentiate multiple domains of sepsis pathobiology, including inflammatory, microvascular, and metabolic dysfunction.

Our findings have several clinical and translational implications. First, they suggest that undernutrition in adults with severe infection should not be viewed solely as a marker of chronic vulnerability or frailty. Rather, severe undernutrition appears to identify a host state characterized by inflammatory, endothelial, and metabolic perturbations that may indicate diminished immune and physiologic resilience during severe infection. Second, our results advance understanding of why undernourished adults experience particularly poor outcomes in LMIC settings where HIV, TB, and other chronic infectious exposures are common, as these intersecting exposures may further compromise host resilience. Third, they highlight the need to improve nutritional assessment and support for adults with severe infection in LMICs, where this aspect of critical care has received far less attention than in HIC settings. Finally, they raise the possibility that host-directed therapeutic strategies in this population may need to account not only for pathogen-specific diagnoses, but also for underlying nutritional state. Our findings also extend the literature on nutrition and critical illness beyond the obesity-focused paradigm that has predominated in HIC settings. Although obesity in sepsis has been associated in some studies with improved survival (the “obesity paradox”), with adipose tissue exerting complex effects on inflammatory host responses (14,15), we observed a contrasting pattern in severe undernutrition. Severe undernutrition was associated with greater physiological instability, higher mortality, and a proteomic profile consistent with inflammatory activation, vascular and tissue stress, and impaired systemic homeostasis, suggesting that severe undernutrition may confer a state of diminished host resilience during severe infection.

Our study has limitations. First, given the observational nature of our cohorts, we cannot determine causal relationships between nutritional status, circulating proteins, and clinical outcomes. Second, although MUAC is a pragmatic, widely utilized measure of nutritional status in LMICs, no guideline-based adult MUAC cutoff exists, and our definitions of severe and mild-moderate undernutrition were based on literature-informed thresholds. Third, our proteomic analysis included 180 circulating proteins and therefore did not capture the full complexity of host-response dysregulation. Fourth, although we adjusted for major covariables including HIV, malaria, and TB infection, residual confounding from unmeasured co-infections (e.g., helminths), micronutrient deficiencies, enteropathy, or other exposures remains possible. Finally, because we did not include uninfected controls, we cannot determine the extent to which the observed proteomic profile reflects undernutrition itself, severe infection, or their interaction.

## Conclusions

Severe undernutrition in adults hospitalized with severe infection in Uganda was associated with greater physiologic instability, higher mortality, and a distinct proteomic profile independent of major coinfections and other key host factors. These findings suggest that severe undernutrition is associated with a biologically distinct pattern of host-response dysregulation in severe infection and may have implications for risk stratification and future host-directed therapeutic approaches.

## Data Availability

Olink proteomic data from RESERVE-U-2 is available on Dryad at https://doi.org/10.5061/dryad.b2rbnzsq2. Additional data is available from the corresponding author upon reasonable request and obtainment of necessary ethical approvals. Analytic R code is available upon request from Drs. Conte Cortez Martins or Cummings.

