## Supplement for "The Impact of Malnutrition on Host Responses to Severe Infection in Adults: A Multicenter Analysis from Uganda"

**Supplemental Tables**

**Table S1: Differential protein expression in patients with severe malnutrition vs. those without malnutrition**. Comparisons adjusted for age, sex, illness duration prior to enrollment, study site (as a fixed effect) and HIV, malaria, and tuberculosis co-infection.

| **Protein** | **logFC** | **AveExpr** | **t** | **P.Value** | **BH-adj.P.Val** |
| --- | --- | --- | --- | --- | --- |
| IGFBP3 | -0.788 | 3.597 | -5.270 | 0.000000219 | 0.0000392 |
| GAS6 | 0.668 | 3.662 | 4.905 | 0.000001340 | 0.0001200 |
| MUC16 | 1.028 | 2.688 | 4.429 | 0.000012100 | 0.0007220 |
| LYVE1 | 0.566 | 5.168 | 4.330 | 0.000018700 | 0.0008350 |
| SERPINA5 | -0.786 | 6.633 | -4.228 | 0.000028900 | 0.0010400 |
| NRP1 | 0.412 | -0.209 | 3.936 | 0.000096800 | 0.0028900 |
| EFEMP1 | 0.527 | 4.992 | 3.824 | 0.000151000 | 0.0038700 |
| FCGR2A | 0.443 | 3.947 | 3.748 | 0.000203000 | 0.0042000 |
| APOM | -0.603 | 4.233 | -3.720 | 0.000226000 | 0.0042000 |
| TNFRSF12A | 0.660 | 6.636 | 3.710 | 0.000235000 | 0.0042000 |
| FETUB | -0.648 | 2.962 | -3.660 | 0.000284000 | 0.0046200 |
| OSMR | 0.289 | 0.501 | 3.625 | 0.000325000 | 0.0048400 |
| FGF2 | 0.515 | 1.290 | 3.588 | 0.000372000 | 0.0051300 |
| PROC | -0.555 | 3.336 | -3.567 | 0.000403000 | 0.0051500 |
| TNFRSF21 | 0.265 | 9.158 | 3.514 | 0.000489000 | 0.0058400 |
| DCN | 0.299 | 2.567 | 3.465 | 0.000586000 | 0.0065500 |
| CAIX | 0.607 | 5.776 | 3.400 | 0.000738000 | 0.0077700 |
| IL-15 | 0.481 | 5.247 | 3.318 | 0.000987000 | 0.0098200 |
| SAA4 | -0.644 | 4.836 | -3.253 | 0.001230000 | 0.0116000 |
| MMP7 | 0.300 | 12.642 | 3.234 | 0.001320000 | 0.0118000 |
| LAMP3 | 0.540 | 6.058 | 3.159 | 0.001700000 | 0.0145000 |
| CNDP1 | -0.579 | 2.455 | -3.132 | 0.001860000 | 0.0151000 |
| CXCL12 | 0.261 | 2.075 | 3.101 | 0.002060000 | 0.0160000 |
| F11 | -0.463 | 6.222 | -2.969 | 0.003160000 | 0.0235000 |
| TIMP1 | 0.427 | 6.480 | 2.907 | 0.003840000 | 0.0268000 |
| SOD1 | 0.417 | -0.892 | 2.903 | 0.003890000 | 0.0268000 |
| CCL20 | 0.783 | 9.785 | 2.808 | 0.005220000 | 0.0336000 |
| IL-6 | 1.006 | 6.277 | 2.805 | 0.005260000 | 0.0336000 |
| CR2 | -0.450 | 5.443 | -2.676 | 0.007740000 | 0.0478000 |
| ICAM1 | 0.341 | 5.717 | 2.640 | 0.008590000 | 0.0512000 |
| F7 | -0.327 | 1.879 | -2.583 | 0.010100000 | 0.0585000 |
| ANGPT2 | 0.328 | 6.170 | 2.529 | 0.011800000 | 0.0659000 |
| TGFBR3 | 0.303 | 2.346 | 2.511 | 0.012400000 | 0.0673000 |
| HGF | 0.393 | 10.285 | 2.486 | 0.013300000 | 0.0701000 |
| TGFBI | 0.319 | 7.623 | 2.441 | 0.015000000 | 0.0769000 |
| VEGFA | 0.296 | 11.104 | 2.351 | 0.019200000 | 0.0953000 |
| REG1A | 0.606 | 7.079 | 2.291 | 0.022500000 | 0.1060000 |
| PRSS2 | 0.638 | 4.549 | 2.289 | 0.022600000 | 0.1060000 |
| CDH1 | 0.283 | 2.313 | 2.262 | 0.024200000 | 0.1110000 |
| PGF | 0.308 | 9.431 | 2.201 | 0.028300000 | 0.1270000 |
| CA1 | 0.500 | 4.937 | 2.190 | 0.029100000 | 0.1270000 |
| SPARCL1 | 0.251 | 2.442 | 2.145 | 0.032500000 | 0.1380000 |
| VASN | 0.194 | 0.901 | 2.131 | 0.033700000 | 0.1400000 |
| PTN | 0.243 | 1.497 | 2.088 | 0.037400000 | 0.1520000 |
| TNC | 0.384 | 4.483 | 2.073 | 0.038800000 | 0.1530000 |
| IL-12 | -0.458 | 6.817 | -2.067 | 0.039300000 | 0.1530000 |
| IGLC2 | 0.204 | 7.031 | 2.030 | 0.043000000 | 0.1640000 |
| MCP1 | 0.375 | 12.070 | 1.985 | 0.047800000 | 0.1780000 |
| PCOLCE | -0.252 | 5.021 | -1.953 | 0.051400000 | 0.1840000 |
| CFHR5 | -0.339 | 7.519 | -1.950 | 0.051900000 | 0.1840000 |
| CX3CL1 | 0.301 | 5.057 | 1.946 | 0.052300000 | 0.1840000 |
| CSF1 | 0.102 | 10.301 | 1.924 | 0.055000000 | 0.1890000 |
| REG3A | 0.260 | 0.606 | 1.910 | 0.056800000 | 0.1920000 |
| CD46 | 0.228 | 2.108 | 1.899 | 0.058200000 | 0.1930000 |
| GAL9 | 0.166 | 9.436 | 1.873 | 0.061700000 | 0.2010000 |
| LILRB5 | 0.246 | 4.925 | 1.762 | 0.078800000 | 0.2520000 |
| ENG | 0.145 | 1.960 | 1.746 | 0.081500000 | 0.2560000 |
| CXCL13 | -0.316 | 9.480 | -1.732 | 0.084100000 | 0.2560000 |
| ANG | -0.236 | 6.108 | -1.726 | 0.085100000 | 0.2560000 |
| TIE2 | 0.138 | 8.084 | 1.723 | 0.085700000 | 0.2560000 |
| LCN2 | 0.297 | 2.595 | 1.713 | 0.087500000 | 0.2570000 |
| VCAM1 | 0.205 | 4.308 | 1.672 | 0.095300000 | 0.2740000 |
| C2 | -0.200 | 4.943 | -1.667 | 0.096300000 | 0.2740000 |
| PD-L2 | 0.180 | 2.516 | 1.638 | 0.102000000 | 0.2860000 |
| DPP4 | 0.194 | 3.493 | 1.623 | 0.105000000 | 0.2900000 |
| ADGRG1 | 0.262 | 1.896 | 1.609 | 0.108000000 | 0.2940000 |
| UMOD | 0.103 | 0.523 | 1.583 | 0.114000000 | 0.3040000 |
| EGF | -0.313 | 10.033 | -1.568 | 0.118000000 | 0.3040000 |
| IL13 | -0.242 | 1.523 | -1.564 | 0.119000000 | 0.3040000 |
| IL-18 | 0.326 | 10.980 | 1.559 | 0.120000000 | 0.3040000 |
| MET | 0.125 | 1.166 | 1.555 | 0.121000000 | 0.3040000 |
| SERPINA7 | -0.178 | 4.082 | -1.537 | 0.125000000 | 0.3110000 |
| TIMD4 | 0.264 | 3.760 | 1.528 | 0.127000000 | 0.3120000 |
| PLTP | 0.131 | 1.365 | 1.510 | 0.132000000 | 0.3190000 |
| TNF | -0.364 | 5.933 | -1.480 | 0.140000000 | 0.3330000 |
| PDCD1 | -0.245 | 8.641 | -1.464 | 0.144000000 | 0.3390000 |
| TNFRSF9 | -0.272 | 7.053 | -1.458 | 0.146000000 | 0.3390000 |
| MCP3 | 0.355 | 3.457 | 1.412 | 0.159000000 | 0.3640000 |
| ARG1 | 0.254 | 3.899 | 1.387 | 0.166000000 | 0.3760000 |
| TNXB | 0.124 | 0.811 | 1.376 | 0.169000000 | 0.3790000 |
| DEFA1 | 0.300 | 1.149 | 1.356 | 0.176000000 | 0.3890000 |
| CXCL11 | 0.379 | 9.691 | 1.315 | 0.189000000 | 0.4110000 |
| CXCL9 | -0.319 | 9.085 | -1.311 | 0.191000000 | 0.4110000 |
| FCGR3B | 0.166 | 3.561 | 1.271 | 0.204000000 | 0.4350000 |
| CD59 | 0.170 | 0.026 | 1.263 | 0.207000000 | 0.4370000 |
| ITGAM | -0.133 | -0.515 | -1.235 | 0.217000000 | 0.4520000 |
| CCL3 | -0.409 | 9.469 | -1.229 | 0.220000000 | 0.4520000 |
| CCL5 | -0.237 | 6.056 | -1.173 | 0.242000000 | 0.4910000 |
| COMP | 0.201 | 6.707 | 1.162 | 0.246000000 | 0.4910000 |
| HO1 | -0.133 | 12.647 | -1.150 | 0.251000000 | 0.4910000 |
| TIE1 | 0.126 | 1.114 | 1.146 | 0.252000000 | 0.4910000 |
| CHL1 | 0.121 | 2.384 | 1.146 | 0.253000000 | 0.4910000 |
| GZMH | 0.260 | 5.794 | 1.122 | 0.262000000 | 0.5040000 |
| MFAP5 | -0.082 | 0.818 | -1.117 | 0.265000000 | 0.5040000 |
| CCL19 | 0.287 | 11.432 | 1.102 | 0.271000000 | 0.5110000 |
| CXCL10 | 0.266 | 10.109 | 1.047 | 0.296000000 | 0.5510000 |
| PRCP | 0.134 | -0.356 | 1.037 | 0.300000000 | 0.5540000 |
| ICAM3 | 0.127 | 2.384 | 0.999 | 0.318000000 | 0.5760000 |
| IL-10 | -0.335 | 6.120 | -0.999 | 0.319000000 | 0.5760000 |
| CRTAC1 | 0.118 | 2.563 | 0.973 | 0.331000000 | 0.5930000 |
| GP1BA | 0.103 | 5.471 | 0.954 | 0.340000000 | 0.5960000 |
| PTPRS | 0.054 | 0.902 | 0.951 | 0.342000000 | 0.5960000 |
| COL18A1 | -0.128 | 2.856 | -0.948 | 0.343000000 | 0.5960000 |
| CD40 | 0.121 | 10.950 | 0.943 | 0.346000000 | 0.5960000 |
| NCR1 | -0.136 | 3.958 | -0.925 | 0.355000000 | 0.6060000 |
| GZMA | -0.145 | 7.558 | -0.914 | 0.361000000 | 0.6100000 |
| ADA | 0.152 | 5.769 | 0.884 | 0.377000000 | 0.6310000 |
| LAG3 | -0.183 | 5.384 | -0.849 | 0.396000000 | 0.6520000 |
| NOS3 | 0.097 | 2.229 | 0.841 | 0.401000000 | 0.6520000 |
| CCL14 | 0.112 | 5.392 | 0.836 | 0.404000000 | 0.6520000 |
| CRTAM | 0.126 | 7.098 | 0.830 | 0.407000000 | 0.6520000 |
| IGFBP6 | -0.121 | 4.980 | -0.825 | 0.410000000 | 0.6520000 |
| CD4 | 0.109 | 4.703 | 0.822 | 0.411000000 | 0.6520000 |
| MEGF9 | 0.076 | 4.034 | 0.807 | 0.420000000 | 0.6600000 |
| NOTCH1 | 0.072 | 3.304 | 0.792 | 0.429000000 | 0.6620000 |
| AOC3 | 0.069 | 1.778 | 0.791 | 0.429000000 | 0.6620000 |
| KIR3DL1 | -0.197 | 3.479 | -0.768 | 0.443000000 | 0.6650000 |
| KLRD1 | 0.134 | 8.574 | 0.766 | 0.444000000 | 0.6650000 |
| IL4 | -0.085 | 1.743 | -0.763 | 0.446000000 | 0.6650000 |
| LILRB2 | 0.120 | 3.629 | 0.763 | 0.446000000 | 0.6650000 |
| CA3 | 0.143 | 1.214 | 0.747 | 0.456000000 | 0.6740000 |
| QPCT | -0.072 | -0.718 | -0.729 | 0.466000000 | 0.6840000 |
| C1QTNF1 | 0.087 | 3.646 | 0.674 | 0.501000000 | 0.7230000 |
| TRAIL | -0.091 | 8.591 | -0.673 | 0.501000000 | 0.7230000 |
| CD28 | 0.071 | 0.988 | 0.663 | 0.507000000 | 0.7260000 |
| ANGPTL3 | -0.090 | 4.937 | -0.657 | 0.511000000 | 0.7260000 |
| PD-L1 | 0.099 | 7.017 | 0.651 | 0.515000000 | 0.7260000 |
| MBL2 | -0.168 | 8.327 | -0.637 | 0.525000000 | 0.7340000 |
| ST6GAL1 | 0.084 | 2.144 | 0.624 | 0.533000000 | 0.7400000 |
| MMP12 | 0.133 | 7.343 | 0.616 | 0.538000000 | 0.7410000 |
| PLXNB2 | 0.055 | 0.645 | 0.596 | 0.551000000 | 0.7530000 |
| LILRB1 | 0.076 | 2.361 | 0.585 | 0.559000000 | 0.7570000 |
| MCP2 | -0.126 | 10.096 | -0.579 | 0.563000000 | 0.7570000 |
| TNFSF14 | -0.096 | 7.177 | -0.574 | 0.566000000 | 0.7570000 |
| CES1 | -0.100 | 1.982 | -0.560 | 0.576000000 | 0.7630000 |
| CD40L | -0.146 | 6.965 | -0.539 | 0.590000000 | 0.7690000 |
| FCN2 | -0.084 | 5.251 | -0.517 | 0.605000000 | 0.7690000 |
| NCAM1 | 0.052 | 1.855 | 0.517 | 0.605000000 | 0.7690000 |
| IL-5 | 0.110 | 1.430 | 0.517 | 0.605000000 | 0.7690000 |
| CCL4 | -0.124 | 9.441 | -0.514 | 0.607000000 | 0.7690000 |
| CCL23 | 0.073 | 11.483 | 0.514 | 0.608000000 | 0.7690000 |
| CD70 | -0.085 | 4.476 | -0.507 | 0.612000000 | 0.7690000 |
| CASP8 | 0.096 | 7.148 | 0.505 | 0.614000000 | 0.7690000 |
| CD5 | 0.061 | 6.444 | 0.491 | 0.623000000 | 0.7720000 |
| NID1 | -0.058 | 3.967 | -0.489 | 0.625000000 | 0.7720000 |
| CXCL1 | 0.055 | 10.881 | 0.478 | 0.633000000 | 0.7740000 |
| GNLY | -0.068 | 1.154 | -0.474 | 0.635000000 | 0.7740000 |
| PAM | 0.042 | 0.657 | 0.454 | 0.650000000 | 0.7860000 |
| VEGFR2 | -0.041 | 7.874 | -0.441 | 0.659000000 | 0.7920000 |
| SELL | 0.042 | 8.985 | 0.402 | 0.688000000 | 0.8210000 |
| LAPTGFB1 | 0.042 | 9.863 | 0.391 | 0.696000000 | 0.8250000 |
| GAL1 | 0.035 | 6.553 | 0.380 | 0.704000000 | 0.8290000 |
| IL2 | -0.027 | 1.385 | -0.372 | 0.710000000 | 0.8300000 |
| TNFRSF4 | 0.059 | 6.363 | 0.362 | 0.718000000 | 0.8320000 |
| CCL18 | -0.070 | 7.949 | -0.356 | 0.722000000 | 0.8320000 |
| CD8A | 0.090 | 9.901 | 0.352 | 0.725000000 | 0.8320000 |
| CD27 | 0.035 | 9.924 | 0.343 | 0.732000000 | 0.8340000 |
| IL-12RB1 | 0.045 | 3.570 | 0.327 | 0.744000000 | 0.8430000 |
| MCP4 | -0.066 | 11.601 | -0.317 | 0.751000000 | 0.8450000 |
| FASLG | -0.048 | 7.277 | -0.312 | 0.755000000 | 0.8450000 |
| CST3 | -0.049 | 6.317 | -0.294 | 0.769000000 | 0.8550000 |
| IFN-γ | -0.103 | 8.724 | -0.249 | 0.804000000 | 0.8880000 |
| PDGFB | 0.025 | 10.797 | 0.226 | 0.821000000 | 0.9020000 |
| IL-7R | 0.021 | 1.452 | 0.201 | 0.840000000 | 0.9170000 |
| CD83 | 0.028 | 3.414 | 0.194 | 0.846000000 | 0.9180000 |
| ICOSLG | -0.019 | 5.574 | -0.176 | 0.861000000 | 0.9280000 |
| ANGPT1 | -0.020 | 9.465 | -0.159 | 0.874000000 | 0.9370000 |
| MICAB | 0.049 | 4.789 | 0.152 | 0.879000000 | 0.9370000 |
| PLA2G7 | -0.011 | 0.967 | -0.123 | 0.902000000 | 0.9540000 |
| THBS4 | 0.022 | 3.513 | 0.118 | 0.906000000 | 0.9540000 |
| CXCL5 | -0.019 | 12.526 | -0.111 | 0.912000000 | 0.9540000 |
| CD244 | 0.011 | 6.451 | 0.092 | 0.927000000 | 0.9620000 |
| IL-7 | -0.016 | 6.070 | -0.088 | 0.930000000 | 0.9620000 |
| GZMB | -0.020 | 3.807 | -0.076 | 0.940000000 | 0.9670000 |
| KIT | 0.007 | 3.505 | 0.066 | 0.947000000 | 0.9690000 |
| TCN2 | 0.006 | 4.470 | 0.043 | 0.966000000 | 0.9820000 |
| CCL17 | -0.007 | 10.733 | -0.033 | 0.974000000 | 0.9820000 |
| CA4 | 0.003 | 1.097 | 0.029 | 0.977000000 | 0.9820000 |
| TWEAK | -0.003 | 9.129 | -0.020 | 0.984000000 | 0.9840000 |

**Table S2: Differential protein expression in patients with severe malnutrition vs. those with mild-to-moderate malnutrition**. Comparisons adjusted for age, sex, illness duration prior to enrollment, study site (as a fixed effect) and HIV, malaria, and tuberculosis co-infection.

| **Protein** | **logFC** | **AveExpr** | **t** | **P.Value** | **BH-adj.P.Val** |
| --- | --- | --- | --- | --- | --- |
| LYVE1 | 0.669 | 5.168 | 5.108 | 0.000000496 | 0.0000887 |
| GAS6 | 0.598 | 3.662 | 4.383 | 0.000014800 | 0.0013200 |
| IGFBP3 | -0.587 | 3.597 | -3.921 | 0.000103000 | 0.0061500 |
| CAIX | 0.665 | 5.776 | 3.722 | 0.000225000 | 0.0093700 |
| NRP1 | 0.386 | -0.209 | 3.681 | 0.000262000 | 0.0093700 |
| CRTAC1 | 0.443 | 2.563 | 3.633 | 0.000314000 | 0.0093700 |
| OSMR | 0.280 | 0.501 | 3.509 | 0.000499000 | 0.0128000 |
| PRCP | 0.448 | -0.356 | 3.457 | 0.000601000 | 0.0131000 |
| SERPINA5 | -0.633 | 6.633 | -3.402 | 0.000733000 | 0.0131000 |
| FGF2 | 0.489 | 1.290 | 3.395 | 0.000752000 | 0.0131000 |
| SOD1 | 0.485 | -0.892 | 3.369 | 0.000823000 | 0.0131000 |
| TNFRSF21 | 0.253 | 9.158 | 3.350 | 0.000880000 | 0.0131000 |
| SAA4 | -0.660 | 4.836 | -3.328 | 0.000952000 | 0.0131000 |
| EFEMP1 | 0.438 | 4.992 | 3.176 | 0.001600000 | 0.0205000 |
| CXCL12 | 0.257 | 2.075 | 3.041 | 0.002510000 | 0.0299000 |
| UMOD | 0.195 | 0.523 | 2.982 | 0.003030000 | 0.0339000 |
| TNFRSF12A | 0.521 | 6.636 | 2.927 | 0.003610000 | 0.0380000 |
| DPP4 | 0.347 | 3.493 | 2.903 | 0.003890000 | 0.0387000 |
| CA1 | 0.650 | 4.937 | 2.843 | 0.004690000 | 0.0442000 |
| DCN | 0.241 | 2.567 | 2.785 | 0.005600000 | 0.0489000 |
| IL-15 | 0.404 | 5.247 | 2.777 | 0.005740000 | 0.0489000 |
| PLTP | 0.239 | 1.365 | 2.761 | 0.006010000 | 0.0489000 |
| FCGR2A | 0.308 | 3.947 | 2.600 | 0.009660000 | 0.0752000 |
| CHL1 | 0.267 | 2.384 | 2.523 | 0.012000000 | 0.0854000 |
| C2 | -0.301 | 4.943 | -2.505 | 0.012600000 | 0.0854000 |
| FETUB | -0.443 | 2.962 | -2.502 | 0.012700000 | 0.0854000 |
| SPARCL1 | 0.292 | 2.442 | 2.491 | 0.013100000 | 0.0854000 |
| F11 | -0.388 | 6.222 | -2.484 | 0.013400000 | 0.0854000 |
| LAG3 | -0.532 | 5.384 | -2.468 | 0.014000000 | 0.0863000 |
| MMP7 | 0.227 | 12.642 | 2.439 | 0.015100000 | 0.0880000 |
| CDH1 | 0.306 | 2.313 | 2.436 | 0.015200000 | 0.0880000 |
| TGFBR3 | 0.291 | 2.346 | 2.414 | 0.016200000 | 0.0908000 |
| CXCL13 | -0.423 | 9.480 | -2.313 | 0.021200000 | 0.1120000 |
| MUC16 | 0.533 | 2.688 | 2.293 | 0.022300000 | 0.1120000 |
| IL-12 | -0.507 | 6.817 | -2.285 | 0.022800000 | 0.1120000 |
| PROC | -0.356 | 3.336 | -2.280 | 0.023100000 | 0.1120000 |
| VASN | 0.208 | 0.901 | 2.278 | 0.023200000 | 0.1120000 |
| HGF | 0.334 | 10.285 | 2.110 | 0.035400000 | 0.1590000 |
| REG1A | 0.554 | 7.079 | 2.090 | 0.037200000 | 0.1590000 |
| APOM | -0.340 | 4.233 | -2.089 | 0.037300000 | 0.1590000 |
| MET | 0.167 | 1.166 | 2.081 | 0.038000000 | 0.1590000 |
| ICAM1 | 0.269 | 5.717 | 2.075 | 0.038600000 | 0.1590000 |
| COMP | 0.359 | 6.707 | 2.072 | 0.038900000 | 0.1590000 |
| TNXB | 0.187 | 0.811 | 2.069 | 0.039200000 | 0.1590000 |
| KIT | 0.228 | 3.505 | 2.025 | 0.043500000 | 0.1730000 |
| TIMP1 | 0.296 | 6.480 | 2.012 | 0.044900000 | 0.1750000 |
| CX3CL1 | 0.310 | 5.057 | 2.002 | 0.045900000 | 0.1750000 |
| REG3A | 0.270 | 0.606 | 1.978 | 0.048500000 | 0.1790000 |
| PDCD1 | -0.330 | 8.641 | -1.973 | 0.049100000 | 0.1790000 |
| TNFRSF9 | -0.362 | 7.053 | -1.940 | 0.053100000 | 0.1900000 |
| CCL20 | 0.511 | 9.785 | 1.830 | 0.068000000 | 0.2350000 |
| CXCL9 | -0.446 | 9.085 | -1.828 | 0.068300000 | 0.2350000 |
| PGF | 0.252 | 9.431 | 1.796 | 0.073200000 | 0.2430000 |
| TGFBI | 0.235 | 7.623 | 1.792 | 0.073900000 | 0.2430000 |
| IL-6 | 0.642 | 6.277 | 1.788 | 0.074500000 | 0.2430000 |
| CA3 | 0.339 | 1.214 | 1.771 | 0.077300000 | 0.2470000 |
| LCN2 | 0.304 | 2.595 | 1.753 | 0.080300000 | 0.2510000 |
| PCOLCE | -0.226 | 5.021 | -1.747 | 0.081400000 | 0.2510000 |
| ENG | 0.143 | 1.960 | 1.717 | 0.086600000 | 0.2630000 |
| CNDP1 | -0.315 | 2.455 | -1.703 | 0.089300000 | 0.2660000 |
| VEGFA | 0.212 | 11.104 | 1.680 | 0.093700000 | 0.2750000 |
| CFHR5 | -0.291 | 7.519 | -1.667 | 0.096200000 | 0.2750000 |
| PTPRS | 0.095 | 0.902 | 1.658 | 0.098100000 | 0.2750000 |
| NCAM1 | 0.167 | 1.855 | 1.650 | 0.099600000 | 0.2750000 |
| LILRB5 | 0.230 | 4.925 | 1.649 | 0.099900000 | 0.2750000 |
| F7 | -0.200 | 1.879 | -1.580 | 0.115000000 | 0.3120000 |
| MCP1 | 0.293 | 12.070 | 1.548 | 0.122000000 | 0.3270000 |
| ANGPT2 | 0.195 | 6.170 | 1.501 | 0.134000000 | 0.3530000 |
| NOS3 | 0.172 | 2.229 | 1.481 | 0.139000000 | 0.3620000 |
| ANG | -0.200 | 6.108 | -1.458 | 0.145000000 | 0.3720000 |
| KIR3DL1 | -0.372 | 3.479 | -1.448 | 0.148000000 | 0.3740000 |
| MMP12 | 0.310 | 7.343 | 1.437 | 0.151000000 | 0.3760000 |
| CCL19 | 0.367 | 11.432 | 1.408 | 0.160000000 | 0.3910000 |
| TCN2 | -0.185 | 4.470 | -1.401 | 0.162000000 | 0.3910000 |
| GAL1 | 0.127 | 6.553 | 1.395 | 0.164000000 | 0.3910000 |
| AOC3 | 0.119 | 1.778 | 1.359 | 0.175000000 | 0.4120000 |
| ARG1 | 0.242 | 3.899 | 1.317 | 0.189000000 | 0.4390000 |
| COL18A1 | -0.176 | 2.856 | -1.297 | 0.195000000 | 0.4480000 |
| PRSS2 | 0.355 | 4.549 | 1.272 | 0.204000000 | 0.4620000 |
| TIMD4 | 0.218 | 3.760 | 1.263 | 0.207000000 | 0.4640000 |
| GZMA | -0.193 | 7.558 | -1.213 | 0.226000000 | 0.4970000 |
| GNLY | -0.174 | 1.154 | -1.204 | 0.229000000 | 0.4970000 |
| CR2 | -0.202 | 5.443 | -1.201 | 0.231000000 | 0.4970000 |
| C1QTNF1 | 0.153 | 3.646 | 1.174 | 0.241000000 | 0.5120000 |
| CCL18 | -0.231 | 7.949 | -1.168 | 0.243000000 | 0.5120000 |
| IL2 | 0.082 | 1.385 | 1.154 | 0.249000000 | 0.5190000 |
| TWEAK | 0.154 | 9.129 | 1.145 | 0.253000000 | 0.5200000 |
| TNF | -0.277 | 5.933 | -1.122 | 0.262000000 | 0.5290000 |
| CCL5 | -0.227 | 6.056 | -1.120 | 0.263000000 | 0.5290000 |
| THBS4 | 0.202 | 3.513 | 1.099 | 0.272000000 | 0.5360000 |
| PTN | 0.128 | 1.497 | 1.099 | 0.273000000 | 0.5360000 |
| HO1 | -0.126 | 12.647 | -1.088 | 0.277000000 | 0.5400000 |
| EGF | -0.215 | 10.033 | -1.074 | 0.284000000 | 0.5460000 |
| CD46 | 0.126 | 2.108 | 1.050 | 0.294000000 | 0.5520000 |
| ADA | 0.180 | 5.769 | 1.049 | 0.295000000 | 0.5520000 |
| NOTCH1 | 0.095 | 3.304 | 1.046 | 0.296000000 | 0.5520000 |
| NID1 | -0.120 | 3.967 | -1.014 | 0.311000000 | 0.5700000 |
| CSF1 | 0.054 | 10.301 | 1.012 | 0.312000000 | 0.5700000 |
| TNC | 0.183 | 4.483 | 0.986 | 0.325000000 | 0.5870000 |
| IFN-γ | -0.399 | 8.724 | -0.960 | 0.338000000 | 0.6030000 |
| VCAM1 | 0.118 | 4.308 | 0.955 | 0.340000000 | 0.6030000 |
| CD70 | -0.159 | 4.476 | -0.946 | 0.345000000 | 0.6050000 |
| TIE1 | 0.099 | 1.114 | 0.905 | 0.366000000 | 0.6360000 |
| CD59 | 0.118 | 0.026 | 0.873 | 0.383000000 | 0.6600000 |
| PD-L2 | 0.094 | 2.516 | 0.853 | 0.394000000 | 0.6720000 |
| MCP2 | -0.176 | 10.096 | -0.810 | 0.418000000 | 0.7060000 |
| SERPINA7 | -0.093 | 4.082 | -0.797 | 0.426000000 | 0.7090000 |
| LAMP3 | 0.136 | 6.058 | 0.794 | 0.428000000 | 0.7090000 |
| LILRB1 | 0.102 | 2.361 | 0.777 | 0.438000000 | 0.7190000 |
| DEFA1 | 0.168 | 1.149 | 0.757 | 0.450000000 | 0.7310000 |
| CD40L | 0.199 | 6.965 | 0.732 | 0.465000000 | 0.7470000 |
| MEGF9 | 0.069 | 4.034 | 0.728 | 0.467000000 | 0.7470000 |
| NCR1 | -0.105 | 3.958 | -0.714 | 0.476000000 | 0.7540000 |
| IL-5 | 0.151 | 1.430 | 0.706 | 0.481000000 | 0.7550000 |
| FCGR3B | 0.087 | 3.561 | 0.667 | 0.505000000 | 0.7830000 |
| IL-7R | 0.068 | 1.452 | 0.663 | 0.507000000 | 0.7830000 |
| CD28 | -0.069 | 0.988 | -0.645 | 0.519000000 | 0.7870000 |
| CD27 | -0.065 | 9.924 | -0.633 | 0.527000000 | 0.7870000 |
| IL13 | -0.098 | 1.523 | -0.630 | 0.529000000 | 0.7870000 |
| CD8A | 0.161 | 9.901 | 0.627 | 0.531000000 | 0.7870000 |
| ADGRG1 | 0.101 | 1.896 | 0.621 | 0.535000000 | 0.7870000 |
| CCL3 | -0.206 | 9.469 | -0.618 | 0.537000000 | 0.7870000 |
| CXCL5 | 0.104 | 12.526 | 0.606 | 0.545000000 | 0.7930000 |
| CD5 | -0.075 | 6.444 | -0.598 | 0.550000000 | 0.7940000 |
| IL-7 | -0.108 | 6.070 | -0.591 | 0.555000000 | 0.7950000 |
| CCL14 | 0.074 | 5.392 | 0.556 | 0.579000000 | 0.8220000 |
| CST3 | -0.090 | 6.317 | -0.541 | 0.589000000 | 0.8280000 |
| IL-18 | 0.111 | 10.980 | 0.531 | 0.596000000 | 0.8280000 |
| PLXNB2 | 0.048 | 0.645 | 0.527 | 0.598000000 | 0.8280000 |
| CCL23 | 0.074 | 11.483 | 0.522 | 0.602000000 | 0.8280000 |
| CES1 | -0.092 | 1.982 | -0.516 | 0.606000000 | 0.8280000 |
| TIE2 | 0.037 | 8.084 | 0.464 | 0.643000000 | 0.8680000 |
| CXCL1 | 0.052 | 10.881 | 0.455 | 0.650000000 | 0.8680000 |
| CCL17 | 0.098 | 10.733 | 0.441 | 0.659000000 | 0.8680000 |
| IGFBP6 | -0.065 | 4.980 | -0.441 | 0.660000000 | 0.8680000 |
| GZMH | 0.102 | 5.794 | 0.441 | 0.660000000 | 0.8680000 |
| CASP8 | 0.081 | 7.148 | 0.424 | 0.672000000 | 0.8780000 |
| GP1BA | 0.044 | 5.471 | 0.406 | 0.685000000 | 0.8850000 |
| ST6GAL1 | -0.055 | 2.144 | -0.403 | 0.687000000 | 0.8850000 |
| PLA2G7 | 0.034 | 0.967 | 0.369 | 0.712000000 | 0.9100000 |
| MCP4 | 0.075 | 11.601 | 0.361 | 0.719000000 | 0.9120000 |
| PDGFB | -0.038 | 10.797 | -0.341 | 0.734000000 | 0.9250000 |
| MCP3 | 0.080 | 3.457 | 0.320 | 0.749000000 | 0.9380000 |
| CD40 | 0.039 | 10.950 | 0.303 | 0.762000000 | 0.9470000 |
| CA4 | 0.028 | 1.097 | 0.277 | 0.782000000 | 0.9480000 |
| TRAIL | -0.036 | 8.591 | -0.268 | 0.789000000 | 0.9480000 |
| QPCT | 0.026 | -0.718 | 0.263 | 0.793000000 | 0.9480000 |
| ANGPTL3 | -0.036 | 4.937 | -0.261 | 0.794000000 | 0.9480000 |
| GAL9 | 0.023 | 9.436 | 0.257 | 0.797000000 | 0.9480000 |
| MFAP5 | 0.019 | 0.818 | 0.255 | 0.799000000 | 0.9480000 |
| KLRD1 | -0.044 | 8.574 | -0.254 | 0.800000000 | 0.9480000 |
| CXCL11 | 0.069 | 9.691 | 0.239 | 0.811000000 | 0.9550000 |
| LILRB2 | 0.036 | 3.629 | 0.228 | 0.820000000 | 0.9580000 |
| ICAM3 | -0.028 | 2.384 | -0.222 | 0.824000000 | 0.9580000 |
| SELL | 0.022 | 8.985 | 0.207 | 0.836000000 | 0.9660000 |
| TNFSF14 | -0.033 | 7.177 | -0.199 | 0.842000000 | 0.9660000 |
| PD-L1 | 0.027 | 7.017 | 0.179 | 0.858000000 | 0.9720000 |
| IL-12RB1 | -0.025 | 3.570 | -0.179 | 0.858000000 | 0.9720000 |
| IL-10 | -0.051 | 6.120 | -0.151 | 0.880000000 | 0.9760000 |
| MBL2 | 0.039 | 8.327 | 0.149 | 0.882000000 | 0.9760000 |
| FASLG | -0.022 | 7.277 | -0.141 | 0.888000000 | 0.9760000 |
| ANGPT1 | 0.017 | 9.465 | 0.136 | 0.892000000 | 0.9760000 |
| MICAB | 0.040 | 4.789 | 0.124 | 0.901000000 | 0.9760000 |
| CCL4 | 0.025 | 9.441 | 0.105 | 0.916000000 | 0.9760000 |
| TNFRSF4 | -0.016 | 6.363 | -0.095 | 0.924000000 | 0.9760000 |
| LAPTGFB1 | 0.010 | 9.863 | 0.095 | 0.924000000 | 0.9760000 |
| CD4 | 0.012 | 4.703 | 0.088 | 0.930000000 | 0.9760000 |
| FCN2 | 0.014 | 5.251 | 0.085 | 0.932000000 | 0.9760000 |
| GZMB | 0.021 | 3.807 | 0.082 | 0.935000000 | 0.9760000 |
| CRTAM | -0.012 | 7.098 | -0.078 | 0.938000000 | 0.9760000 |
| ICOSLG | 0.008 | 5.574 | 0.074 | 0.941000000 | 0.9760000 |
| IL4 | -0.008 | 1.743 | -0.072 | 0.942000000 | 0.9760000 |
| CXCL10 | -0.017 | 10.109 | -0.068 | 0.946000000 | 0.9760000 |
| IGLC2 | 0.006 | 7.031 | 0.064 | 0.949000000 | 0.9760000 |
| CD83 | 0.007 | 3.414 | 0.051 | 0.959000000 | 0.9810000 |
| VEGFR2 | -0.004 | 7.874 | -0.045 | 0.964000000 | 0.9810000 |
| CD244 | 0.004 | 6.451 | 0.033 | 0.974000000 | 0.9820000 |
| PAM | -0.003 | 0.657 | -0.030 | 0.976000000 | 0.9820000 |
| ITGAM | 0.000 | -0.515 | 0.002 | 0.999000000 | 0.9990000 |

**Table S3: Differential protein expression in patients with mild-to-moderate malnutrition vs. those without malnutrition.** Comparisons adjusted for age, sex, illness duration prior to enrollment, study site (as a fixed effect) and HIV, malaria, and tuberculosis co-infection.

| **Prot** | **logFC** | **AveExpr** | **t** | **P.Value** | **BH-adj.P.Val** |
| --- | --- | --- | --- | --- | --- |
| CRTAC1 | -0.324 | 2.563 | -4.098 | 0.0000501 | 0.00897 |
| PRCP | -0.314 | -0.356 | -3.728 | 0.0002190 | 0.01880 |
| LAMP3 | 0.404 | 6.058 | 3.633 | 0.0003150 | 0.01880 |
| MUC16 | 0.495 | 2.688 | 3.277 | 0.0011400 | 0.05090 |
| IGLC2 | 0.198 | 7.031 | 3.021 | 0.0026700 | 0.08110 |
| KIT | -0.221 | 3.505 | -3.016 | 0.0027200 | 0.08110 |
| APOM | -0.264 | 4.233 | -2.501 | 0.0128000 | 0.26700 |
| LAG3 | 0.349 | 5.384 | 2.495 | 0.0130000 | 0.26700 |
| GAL9 | 0.143 | 9.436 | 2.483 | 0.0134000 | 0.26700 |
| IL2 | -0.109 | 1.385 | -2.349 | 0.0193000 | 0.34500 |
| CR2 | -0.248 | 5.443 | -2.264 | 0.0241000 | 0.39200 |
| TCN2 | 0.191 | 4.470 | 2.223 | 0.0267000 | 0.39700 |
| CNDP1 | -0.264 | 2.455 | -2.192 | 0.0289000 | 0.39700 |
| UMOD | -0.092 | 0.523 | -2.158 | 0.0315000 | 0.39700 |
| CHL1 | -0.146 | 2.384 | -2.123 | 0.0343000 | 0.39700 |
| MFAP5 | -0.101 | 0.818 | -2.110 | 0.0355000 | 0.39700 |
| IGFBP3 | -0.201 | 3.597 | -2.063 | 0.0397000 | 0.41800 |
| CD28 | 0.140 | 0.988 | 2.012 | 0.0448000 | 0.42300 |
| DPP4 | -0.153 | 3.493 | -1.975 | 0.0489000 | 0.42300 |
| PROC | -0.200 | 3.336 | -1.971 | 0.0493000 | 0.42300 |
| CD40L | -0.345 | 6.965 | -1.955 | 0.0513000 | 0.42300 |
| TIE2 | 0.101 | 8.084 | 1.934 | 0.0538000 | 0.42300 |
| PLTP | -0.108 | 1.365 | -1.929 | 0.0543000 | 0.42300 |
| ITGAM | -0.134 | -0.515 | -1.901 | 0.0580000 | 0.43200 |
| ICAM3 | 0.156 | 2.384 | 1.878 | 0.0611000 | 0.43800 |
| TWEAK | -0.156 | 9.129 | -1.794 | 0.0735000 | 0.49200 |
| FETUB | -0.204 | 2.962 | -1.774 | 0.0768000 | 0.49200 |
| FCGR2A | 0.135 | 3.947 | 1.759 | 0.0793000 | 0.49200 |
| NCAM1 | -0.115 | 1.855 | -1.746 | 0.0816000 | 0.49200 |
| CXCL10 | 0.284 | 10.109 | 1.714 | 0.0873000 | 0.49200 |
| MCP3 | 0.275 | 3.457 | 1.678 | 0.0940000 | 0.49200 |
| CD5 | 0.136 | 6.444 | 1.676 | 0.0945000 | 0.49200 |
| TNC | 0.201 | 4.483 | 1.668 | 0.0960000 | 0.49200 |
| CXCL11 | 0.310 | 9.691 | 1.653 | 0.0991000 | 0.49200 |
| ST6GAL1 | 0.139 | 2.144 | 1.579 | 0.1150000 | 0.49200 |
| IL-18 | 0.215 | 10.980 | 1.579 | 0.1150000 | 0.49200 |
| CA3 | -0.196 | 1.214 | -1.579 | 0.1150000 | 0.49200 |
| ANGPT2 | 0.133 | 6.170 | 1.577 | 0.1160000 | 0.49200 |
| KLRD1 | 0.178 | 8.574 | 1.569 | 0.1180000 | 0.49200 |
| GAL1 | -0.093 | 6.553 | -1.563 | 0.1190000 | 0.49200 |
| PRSS2 | 0.283 | 4.549 | 1.559 | 0.1200000 | 0.49200 |
| IL-6 | 0.364 | 6.277 | 1.559 | 0.1200000 | 0.49200 |
| F7 | -0.127 | 1.879 | -1.538 | 0.1250000 | 0.49200 |
| QPCT | -0.098 | -0.718 | -1.525 | 0.1280000 | 0.49200 |
| PTN | 0.115 | 1.497 | 1.519 | 0.1300000 | 0.49200 |
| ADGRG1 | 0.161 | 1.896 | 1.517 | 0.1300000 | 0.49200 |
| THBS4 | -0.180 | 3.513 | -1.511 | 0.1320000 | 0.49200 |
| CD27 | 0.100 | 9.924 | 1.502 | 0.1340000 | 0.49200 |
| CCL20 | 0.272 | 9.785 | 1.499 | 0.1350000 | 0.49200 |
| IL13 | -0.144 | 1.523 | -1.434 | 0.1520000 | 0.54600 |
| COMP | -0.158 | 6.707 | -1.404 | 0.1610000 | 0.55200 |
| CSF1 | 0.048 | 10.301 | 1.399 | 0.1620000 | 0.55200 |
| CRTAM | 0.138 | 7.098 | 1.396 | 0.1640000 | 0.55200 |
| TIMP1 | 0.131 | 6.480 | 1.371 | 0.1710000 | 0.56700 |
| IL-10 | -0.284 | 6.120 | -1.302 | 0.1940000 | 0.61600 |
| CD46 | 0.102 | 2.108 | 1.302 | 0.1940000 | 0.61600 |
| C2 | 0.101 | 4.943 | 1.295 | 0.1960000 | 0.61600 |
| MMP12 | -0.177 | 7.343 | -1.266 | 0.2060000 | 0.63100 |
| SERPINA5 | -0.152 | 6.633 | -1.261 | 0.2080000 | 0.63100 |
| CCL18 | 0.161 | 7.949 | 1.251 | 0.2110000 | 0.63100 |
| MMP7 | 0.073 | 12.642 | 1.215 | 0.2250000 | 0.63900 |
| MBL2 | -0.207 | 8.327 | -1.208 | 0.2280000 | 0.63900 |
| LYVE1 | -0.103 | 5.168 | -1.208 | 0.2280000 | 0.63900 |
| PD-L2 | 0.086 | 2.516 | 1.205 | 0.2290000 | 0.63900 |
| TNFRSF12A | 0.139 | 6.636 | 1.197 | 0.2320000 | 0.63900 |
| SERPINA7 | -0.086 | 4.082 | -1.135 | 0.2570000 | 0.68700 |
| CD4 | 0.098 | 4.703 | 1.128 | 0.2600000 | 0.68700 |
| GNLY | 0.105 | 1.154 | 1.124 | 0.2610000 | 0.68700 |
| CXCL5 | -0.123 | 12.526 | -1.103 | 0.2710000 | 0.68700 |
| VCAM1 | 0.088 | 4.308 | 1.098 | 0.2730000 | 0.68700 |
| IFN-γ | 0.296 | 8.724 | 1.095 | 0.2740000 | 0.68700 |
| PTPRS | -0.041 | 0.902 | -1.090 | 0.2760000 | 0.68700 |
| TNXB | -0.063 | 0.811 | -1.069 | 0.2860000 | 0.68800 |
| IL4 | -0.077 | 1.743 | -1.061 | 0.2890000 | 0.68800 |
| KIR3DL1 | 0.175 | 3.479 | 1.049 | 0.2950000 | 0.68800 |
| GZMH | 0.158 | 5.794 | 1.046 | 0.2960000 | 0.68800 |
| MCP4 | -0.141 | 11.601 | -1.043 | 0.2980000 | 0.68800 |
| DCN | 0.058 | 2.567 | 1.038 | 0.3000000 | 0.68800 |
| VEGFA | 0.084 | 11.104 | 1.027 | 0.3050000 | 0.69100 |
| CA1 | -0.150 | 4.937 | -1.011 | 0.3130000 | 0.69600 |
| TGFBI | 0.085 | 7.623 | 0.994 | 0.3210000 | 0.69600 |
| EFEMP1 | 0.088 | 4.992 | 0.987 | 0.3240000 | 0.69600 |
| NOS3 | -0.074 | 2.229 | -0.987 | 0.3240000 | 0.69600 |
| CD40 | 0.082 | 10.950 | 0.982 | 0.3270000 | 0.69600 |
| CCL4 | -0.149 | 9.441 | -0.953 | 0.3410000 | 0.71900 |
| CCL3 | -0.203 | 9.469 | -0.937 | 0.3490000 | 0.72200 |
| FCGR3B | 0.079 | 3.561 | 0.928 | 0.3540000 | 0.72200 |
| FCN2 | -0.098 | 5.251 | -0.926 | 0.3550000 | 0.72200 |
| DEFA1 | 0.132 | 1.149 | 0.919 | 0.3590000 | 0.72200 |
| CXCL13 | 0.107 | 9.480 | 0.899 | 0.3690000 | 0.73400 |
| AOC3 | -0.050 | 1.778 | -0.877 | 0.3810000 | 0.74700 |
| PDGFB | 0.064 | 10.797 | 0.871 | 0.3840000 | 0.74700 |
| ICAM1 | 0.073 | 5.717 | 0.863 | 0.3890000 | 0.74800 |
| GP1BA | 0.059 | 5.471 | 0.842 | 0.4000000 | 0.74900 |
| IL-15 | 0.078 | 5.247 | 0.824 | 0.4100000 | 0.74900 |
| LILRB2 | 0.084 | 3.629 | 0.821 | 0.4120000 | 0.74900 |
| MET | -0.043 | 1.166 | -0.814 | 0.4160000 | 0.74900 |
| NID1 | 0.062 | 3.967 | 0.810 | 0.4180000 | 0.74900 |
| CXCL9 | 0.127 | 9.085 | 0.799 | 0.4250000 | 0.74900 |
| GAS6 | 0.070 | 3.662 | 0.790 | 0.4300000 | 0.74900 |
| PDCD1 | 0.086 | 8.641 | 0.788 | 0.4310000 | 0.74900 |
| IL-12RB1 | 0.070 | 3.570 | 0.777 | 0.4370000 | 0.74900 |
| IL-7 | 0.092 | 6.070 | 0.774 | 0.4400000 | 0.74900 |
| C1QTNF1 | -0.065 | 3.646 | -0.772 | 0.4410000 | 0.74900 |
| PLA2G7 | -0.045 | 0.967 | -0.758 | 0.4490000 | 0.74900 |
| EGF | -0.098 | 10.033 | -0.757 | 0.4500000 | 0.74900 |
| TNFRSF9 | 0.090 | 7.053 | 0.746 | 0.4560000 | 0.74900 |
| PAM | 0.045 | 0.657 | 0.744 | 0.4580000 | 0.74900 |
| F11 | -0.075 | 6.222 | -0.739 | 0.4600000 | 0.74900 |
| CCL17 | -0.105 | 10.733 | -0.730 | 0.4660000 | 0.74900 |
| SOD1 | -0.068 | -0.892 | -0.726 | 0.4680000 | 0.74900 |
| PD-L1 | 0.072 | 7.017 | 0.725 | 0.4690000 | 0.74900 |
| IL-7R | -0.047 | 1.452 | -0.712 | 0.4770000 | 0.75600 |
| TNFRSF4 | 0.074 | 6.363 | 0.702 | 0.4830000 | 0.75800 |
| CD70 | 0.074 | 4.476 | 0.676 | 0.5000000 | 0.77800 |
| MCP1 | 0.082 | 12.070 | 0.668 | 0.5040000 | 0.77800 |
| TRAIL | -0.055 | 8.591 | -0.623 | 0.5340000 | 0.81100 |
| PGF | 0.056 | 9.431 | 0.617 | 0.5370000 | 0.81100 |
| VEGFR2 | -0.036 | 7.874 | -0.609 | 0.5430000 | 0.81100 |
| ANGPTL3 | -0.054 | 4.937 | -0.608 | 0.5430000 | 0.81100 |
| CD59 | 0.052 | 0.026 | 0.597 | 0.5510000 | 0.81500 |
| IGFBP6 | -0.056 | 4.980 | -0.589 | 0.5560000 | 0.81600 |
| TNFSF14 | -0.062 | 7.177 | -0.575 | 0.5650000 | 0.82000 |
| HGF | 0.059 | 10.285 | 0.572 | 0.5680000 | 0.82000 |
| TNF | -0.088 | 5.933 | -0.547 | 0.5840000 | 0.83400 |
| COL18A1 | 0.047 | 2.856 | 0.539 | 0.5900000 | 0.83400 |
| SPARCL1 | -0.041 | 2.442 | -0.537 | 0.5910000 | 0.83400 |
| CAIX | -0.059 | 5.776 | -0.504 | 0.6150000 | 0.85900 |
| CCL19 | -0.080 | 11.432 | -0.473 | 0.6360000 | 0.88200 |
| GZMA | 0.048 | 7.558 | 0.462 | 0.6440000 | 0.88200 |
| LAPTGFB1 | 0.032 | 9.863 | 0.454 | 0.6500000 | 0.88200 |
| ANGPT1 | -0.037 | 9.465 | -0.454 | 0.6500000 | 0.88200 |
| CFHR5 | -0.049 | 7.519 | -0.430 | 0.6680000 | 0.88600 |
| CCL14 | 0.037 | 5.392 | 0.430 | 0.6680000 | 0.88600 |
| CD8A | -0.071 | 9.901 | -0.424 | 0.6720000 | 0.88600 |
| ANG | -0.036 | 6.108 | -0.407 | 0.6840000 | 0.88600 |
| TIMD4 | 0.045 | 3.760 | 0.404 | 0.6860000 | 0.88600 |
| NOTCH1 | -0.023 | 3.304 | -0.394 | 0.6940000 | 0.88600 |
| ICOSLG | -0.027 | 5.574 | -0.384 | 0.7010000 | 0.88600 |
| NRP1 | 0.026 | -0.209 | 0.383 | 0.7020000 | 0.88600 |
| CA4 | -0.025 | 1.097 | -0.383 | 0.7020000 | 0.88600 |
| CST3 | 0.041 | 6.317 | 0.381 | 0.7030000 | 0.88600 |
| TIE1 | 0.026 | 1.114 | 0.368 | 0.7130000 | 0.89300 |
| MCP2 | 0.051 | 10.096 | 0.358 | 0.7200000 | 0.89600 |
| IL-12 | 0.049 | 6.817 | 0.341 | 0.7330000 | 0.90500 |
| NCR1 | -0.031 | 3.958 | -0.324 | 0.7460000 | 0.91100 |
| PCOLCE | -0.026 | 5.021 | -0.313 | 0.7550000 | 0.91100 |
| REG1A | 0.052 | 7.079 | 0.303 | 0.7620000 | 0.91100 |
| SELL | 0.020 | 8.985 | 0.300 | 0.7640000 | 0.91100 |
| LILRB1 | -0.025 | 2.361 | -0.297 | 0.7670000 | 0.91100 |
| IL-5 | -0.041 | 1.430 | -0.292 | 0.7700000 | 0.91100 |
| FGF2 | 0.027 | 1.290 | 0.288 | 0.7730000 | 0.91100 |
| CDH1 | -0.022 | 2.313 | -0.275 | 0.7840000 | 0.91700 |
| FASLG | -0.026 | 7.277 | -0.263 | 0.7930000 | 0.92200 |
| ADA | -0.029 | 5.769 | -0.255 | 0.7980000 | 0.92200 |
| TNFRSF21 | 0.012 | 9.158 | 0.243 | 0.8080000 | 0.92200 |
| GZMB | -0.041 | 3.807 | -0.242 | 0.8090000 | 0.92200 |
| VASN | -0.014 | 0.901 | -0.233 | 0.8160000 | 0.92400 |
| CD83 | 0.021 | 3.414 | 0.220 | 0.8260000 | 0.93000 |
| OSMR | 0.009 | 0.501 | 0.169 | 0.8660000 | 0.96200 |
| LILRB5 | 0.015 | 4.925 | 0.169 | 0.8660000 | 0.96200 |
| TGFBR3 | 0.011 | 2.346 | 0.144 | 0.8860000 | 0.96900 |
| SAA4 | 0.016 | 4.836 | 0.123 | 0.9020000 | 0.96900 |
| CASP8 | 0.015 | 7.148 | 0.123 | 0.9020000 | 0.96900 |
| MEGF9 | 0.007 | 4.034 | 0.120 | 0.9040000 | 0.96900 |
| REG3A | -0.010 | 0.606 | -0.110 | 0.9130000 | 0.96900 |
| ARG1 | 0.013 | 3.899 | 0.105 | 0.9160000 | 0.96900 |
| PLXNB2 | 0.006 | 0.645 | 0.105 | 0.9160000 | 0.96900 |
| HO1 | -0.007 | 12.647 | -0.093 | 0.9260000 | 0.96900 |
| CD244 | 0.007 | 6.451 | 0.091 | 0.9280000 | 0.96900 |
| CX3CL1 | -0.009 | 5.057 | -0.091 | 0.9280000 | 0.96900 |
| CXCL12 | 0.005 | 2.075 | 0.084 | 0.9330000 | 0.96900 |
| CCL5 | -0.010 | 6.056 | -0.078 | 0.9380000 | 0.96900 |
| LCN2 | -0.008 | 2.595 | -0.066 | 0.9470000 | 0.96900 |
| CES1 | -0.008 | 1.982 | -0.066 | 0.9470000 | 0.96900 |
| MICAB | 0.009 | 4.789 | 0.042 | 0.9660000 | 0.97700 |
| ENG | 0.002 | 1.960 | 0.040 | 0.9680000 | 0.97700 |
| CXCL1 | 0.003 | 10.881 | 0.035 | 0.9720000 | 0.97700 |
| CCL23 | -0.001 | 11.483 | -0.013 | 0.9890000 | 0.98900 |

**Figure S1: Study Flow Diagram**

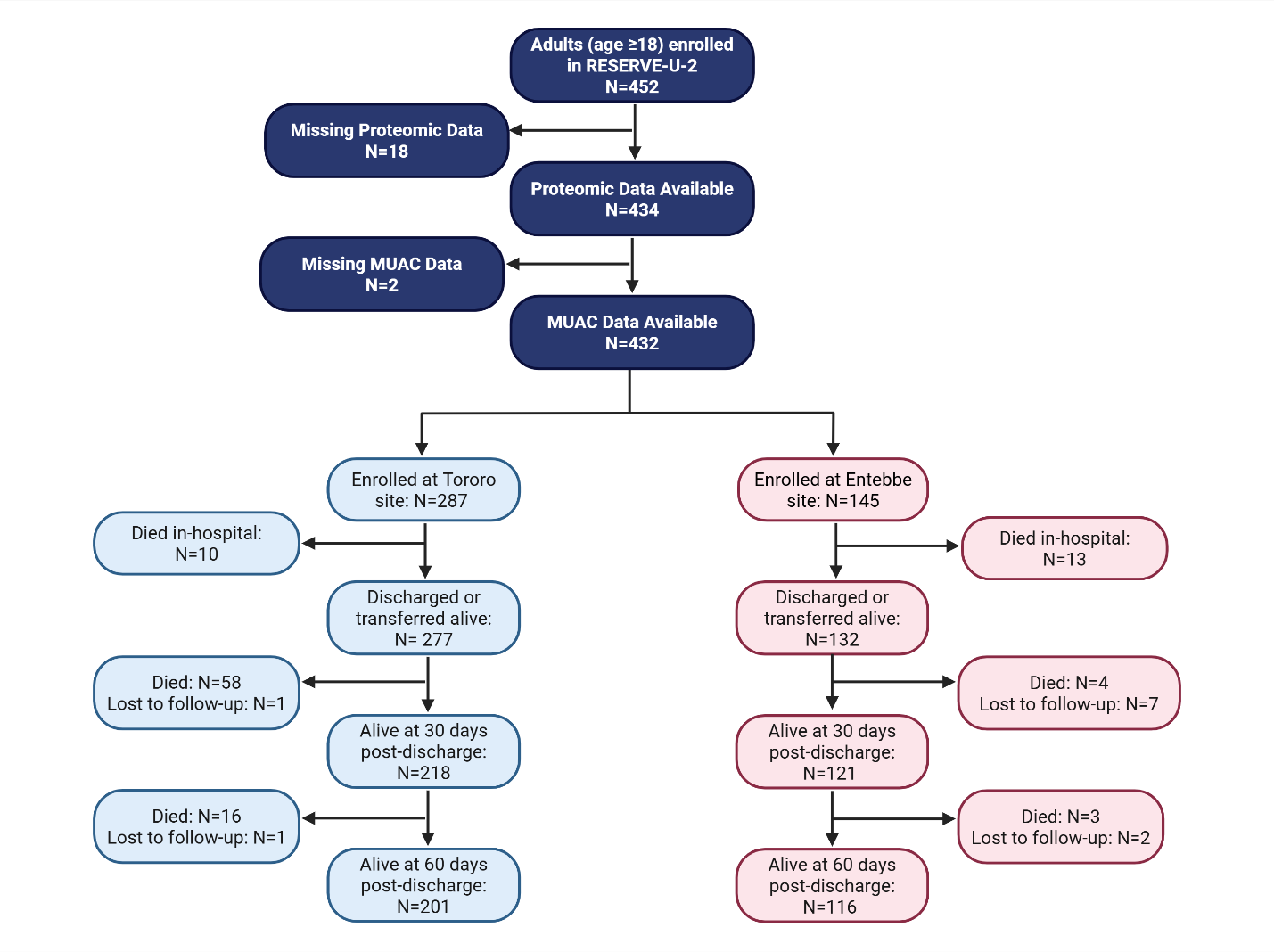
